# Post-Disease Divergence in SARS-CoV-2 RNA Detection between Nasopharyngeal, Anterior Nares and Saliva/Oral Fluid Specimens - Significant Implications for Policy & Public Health

**DOI:** 10.1101/2021.01.26.21250523

**Authors:** Fred Turner, Amy Vandenberg, Vladimir I. Slepnev, Suzana Car, Rita E. Starritt, Michael V. Seger, Noah Kojima, Nina Nirema, Lauren Lopez, Matthew Brobeck, Sarah K. August, Alejandra Orosco, Fred Hertlein, Arthur M. Baca

## Abstract

**Background:** Patients have been shown to shed SARS-CoV-2 viral RNA in nasopharyngeal (NP) specimens for over 100 days after resolution of clinical disease (1, 2). How this relates to anterior nares and oral fluid specimens has not previously been investigated.

**Methods:** We prospectively collected oral fluid, anterior nares, NP swab and serum specimens from 1,326 individuals at 2 “drive-through” testing locations. The Curative SARS-CoV-2 Assay (Curative Assay)(3) on oral fluid and anterior nares specimens was compared to the EURORealTime SARS-CoV-2 Assay (EuroRT Assay)(4) on anterior nares and NP specimens. Viral culture and IgG serology were used to assess infectious potential and stage of disease.

Additionally we investigated differences in viral RNA detection between specimen types, both early (< 21 days) and late (> 21 days) in SARS-CoV-2 infection, by using an employee surveillance program with daily SARS-CoV-2 testing to precisely determine infection date, even without symptoms. We prospectively collected oral fluid, anterior nares and NP swab specimens from 165 subjects with early infections and 22 subjects with late infections. Specimens were tested using the Curative Assay with the “high-sensitivity” Hologic Aptima SARS-CoV-2 Assay (Hologic Assay)(5) on an NP swab used as the comparator. Late infection specimens were also tested with EuroRT and Zymo Quick SARS-CoV-2 rRT-PCR Kit (Zymo) (6) Assays.

**Results:** The “drive-through” study showed similar sensitivities of oral fluid and anterior nares specimens on the Curative Assay to anterior nares specimens tested with the EuroRT Assay. However NP specimens tested with the same EuroRT assay produced 20-30% more positives. Incorporating viral culture and serology data to exclude NP RT-PCR positives that are not infectious or late in the course of disease showed a Positive Percent Agreement (PPA) for of 98.2% and 96.2% and Negative Percent Agreement (NPA) of 97.6% and 98.1% for anterior nares and oral fluid specimens, respectively.

Within 21 days of infection, the Curative Assay showed a PPA and NPA of 100% and 100%, respectively for oral fluid; of 100% and 99% respectively for anterior nares; and of 98.2% and 99.0%, respectively in nasopharyngeal specimens compared to an NP specimen on the Hologic Assay. 29 positives were asymptomatic and showed 100% PPA and 100% NPA for all specimen types. After 21 days from infection onset, significant divergence between NP and other specimen types occurred on all 4 assays. Out of 22 paired sample sets, 18, 13, 8 and 4 NP specimens were positive on the Curative, Zymo, Hologic and EuroRT assays, respectively, compared to only 3, 2, 0 and 1 positive anterior nares specimens. Only one oral fluid sample was positive in both the Curative and Zymo assays.

**Conclusions:** We used a unique population to show significant divergence between NP specimens and anterior nares or oral fluid specimens >21 days from SARS-CoV-2 infection, which appears to be biological variation and is independent of assay used. This has significant public health implications for the use of NP specimens in community testing programs and policy implications for evaluation of novel specimen types and tests where the use of NP swabs as a comparator may say more about the study population than the assay or specimen type to be evaluated and may unnecessarily limit access to testing.

## Introduction

In the 12 months since the first cases of COVID-19, the disease caused by severe acute respiratory syndrome coronavirus 2 (SARS-CoV-2), were identified in Wuhan, China, the outbreak has burgeoned into a global pandemic claiming upwards of 1.8 million lives (7). Identifying and isolating infectious individuals through large scale community testing and contact tracing, at a scale never previously seen, has been critical to containing the spread of the virus (8). Soaring testing demands have paved the way for new modalities of sample collection, such as drive-through(9, 10) and walk-up sites, kiosks, mobile vans(11), and home collection (12).

Real-time reverse transcription-polymerase chain reaction (RT-PCR) testing of nasopharyngeal (NP) specimens has long been considered the “gold standard” of respiratory virus detection and was rapidly adopted for the detection of SARS-CoV-2 (13). However, due to the invasive nature of NP swab collection and the skilled medical staffing required for NP swab collection at scale, many tests from alternative and self-collected sample types have been considered and received Emergency Use Authorization (EUA) by the FDA, such as anterior nares, oral fluid(14) and saliva (15, 16)

SARS-CoV-2 viral load dynamics present a particular challenge for diagnostic testing with some patients continuing to have detectable viral RNA in NP specimens for over 100 days, despite resolution of symptoms (1, 2, 17, 18). Many attempts have been made to isolate replication-competent virus or demonstrate infectious potential through contact tracing of these “persistent-positive” individuals, without success (19, 20, 21, 22). Infectious virus has only been shown to be present for a maximum of 20 days, except in exceptional cases (21). The vast majority of cases harbor infectious virus no more than 8 to 9 days after symptoms first appear leading to current CDC recommendation of release from quarantine 10 days after symptom onset in uncomplicated cases (23).

The effect of this “long-tail” of RNA-positivity on public health outcomes in outpatient testing, frequently being conducted in non-traditional healthcare settings, has not previously been examined. Further, the variation between persistent viral RNA detection after clinical disease between NP specimens and other specimen types such as anterior nares, oral fluid and saliva has not been widely studied.

We hypothesized persistent RNA-positivity occurs to a much greater extent in NP specimens than anterior nares or oral fluid. Thus anterior nares and oral fluid sampling should correlate better with active SARS-CoV-2 infection and transmission risk thus minimizing the inadvertent detection of previously resolved infection. This has profound implications for the design of public health contact tracing programs as well as regulatory policy surrounding how the performance of new tests is studied.

Here we describe the results of a prospective 1,326 patient study comparing NP, anterior nares and oral fluid specimens collected in a real-world “drive-through” setting, incorporating the use of viral culture and serology to assess stage of clinical disease in both symptomatic and asymptomatic individuals. Additionally we describe a study in a highly controlled setting with 187 individuals with months-long history of daily SARS-CoV-2 testing, allowing for precise determination of date of infection, regardless of symptoms, in order to determine the time-course of RNA-positivity in different specimens.

## Materials and Methods

### “Drive through” Study

#### Study Design

We conducted a prospective, specimen comparison study of paired NP, anterior nares and oral fluid specimens collected within one hour from both symptomatic and asymptomatic individuals at two geographically distinct “mass-testing drive-through” sites in Los Angeles, CA and San Antonio, TX. Specimens were tested with 2 different FDA-EUA RT-PCR tests as well as with viral culture and serological testing for anti-SARS-CoV-2 IgG to distinguish early-infectious cases from later and almost certainly noninfectious cases (21) given the lack of available medical history in this setting (24). Symptom status was assessed by a physician.

#### Clinical Sample Collection

Participants provided up to five samples collected within one hour: self-collected oral fluid swab, self-collected anterior nares swab, a provider-collected NP swab, a provider-collected anterior nares sample and a 10 mL venous blood draw.

Subjects were trained on proper self-collection technique and self-collection of the anterior nares and oral fluid specimens occurred without direct supervision or direction. Self-collected oral fluid and anterior nares specimens were placed in specimen collection tubes containing 1 mL of DNA/RNA Shield (Zymo Research, Irvine, CA), a guanidinium-based preservative that deactivates SARS-CoV-2 upon contact, and kept at ambient temperature. Provider-collected NP and anterior nares specimens were collected into 3 mL of Viral Transport Medium (VTM) and refrigerated at 2-8°C to enable viral culture.

#### Specimen Testing

Self-collected oral fluid and anterior nares specimens were tested using the Curative SARS-CoV-2 Multiplex Assay (Curative Assay) (EUA200132/S002/S004). Provider-collected anterior nares and NP specimens were tested using the EURORealTime SARS-CoV-2 Assay (EuroRT Assay) (EUA200525). Serum specimens with the EUROIMMUN Anti-SARS-CoV-2 ELISA (IgG) (EUA200523)(25). Testing was conducted at Curative Labs Inc. (San Dimas, CA) and assays were run per the manufacturer’s instructions for use. Patient identifiers were removed so laboratory personnel were blinded prior to testing. Remnant VTM specimens were stored to enable later evaluation by viral culture. More information is provided in the Supplementary Methods.

#### Viral Culturing

Provider-collected NP and anterior nares specimens for specimens with a Cycle Threshold (Ct) up to 45 were shipped on dry ice to the George Washington University School of Public Health for viral culture analysis. RT-PCR results were not disclosed to the reference culture laboratory. Briefly, samples were serially diluted and incubated with Vero E6 cells which were monitored for cytopathogenic effect. Further protocol details are provided in the Supplementary Methods.

### Early & Late COVID-19 Study

#### Study Design

Subjects were recruited based upon known negative and positive SARS-CoV-2 infection status gathered through routine employee testing protocol using the Curative Assay and assigned to the “early” or “late” group. Both groups were identified as having no known history of SARS-CoV-2 infection where initial infection data was established as the first positive test for prior SARS-CoV-2 infection before the current episode. Individuals with an initial positive test for SARS-CoV-2 within the last 21 days were prospectively assigned to the early group. Individuals with an initial positive test for SARS-CoV-2 infection for more than 22 days and up to 70 days were prospectively assigned to the late group.

#### Early Group Clinical Specimen Collection

All participants provided four specimens: self-collected oral fluid, self-collected anterior nares, and two provider-collected NP specimens, one collected in VTM and one into DNA/RNA Shield. Specimens were collected as previously described for the “Drive through” study, except that self-collected oral fluid and anterior nares samples were collected under the observation and direction of a trained Healthcare Worker (HCW). All samples were collected within a maximum of 24 hours. The collection order of anterior nares and NP specimens was randomly assigned and recorded.

Specimen collection occurred at 4 locations in San Dimas, CA; Torrance, CA; Washington, DC; and Pflugerville, TX with a target enrollment of 60 positive and 100 negative individuals.

#### Late Group Clinical Specimen Collection

Participants provided 5 specimens: self-collected oral fluid swab (DNA/RNA Shield), self-collected anterior nares swab (DNA/RNA Shield), provider-collected anterior nares swab (VTM) and two provider-collected NP swabs (one DNA/RNA Shield, one VTM). Specimens were collected as previously described for the “Drive through” study, except that self-collected oral fluid and anterior nares samples were collected under the observation and direction of a trained Healthcare Worker (HCW). All samples were collected within a maximum of 24 hours. The collection order of anterior nares and NP specimens was randomly assigned and recorded. Oral fluid specimens were collected first so that any effect of the coughing on anterior nares or NP specimens would be equal. Specimen collection occurred only in San Dimas, CA with a target enrollment of 20 individuals.

#### Clinical Specimen Testing

Self-collected specimens were processed at the nearest Curative Labs Inc. clinical laboratory (San Dimas, CA; Washington, DC; or Pflugerville, TX) using the Curative SARS-CoV-2 Assay and remnant specimens from the late group were also analyzed using the Quick SARS-CoV-2 rRT-PCR Kit (Zymo Research Inc., Irvine, CA) which is also authorized for samples collected in DNA/RNA Shield. NP specimens collected in DNA/RNA Shield were processed at Curative Labs Inc. in San Dimas, CA using the Curative SARS-CoV-2 Assay. An aliquot from NP specimens collected in VTM was transferred to the Hologic Specimen Lysis Tube for both early and late groups and was processed by an external and independent CLIA-certified laboratory (BioCollections Worldwide, Inc. Miami, FL) using the Hologic Aptima SARS-CoV-2 Assay (5) on the Hologic Panther System. The additional VTM samples collected in the late group were used to run the EUROIMMUN EuroRealTime SARS-CoV-2 Assay (EUA200525) (4) using the CMG-2017 Prepito Viral DNA/RNA300 Kit (Chemagen), and the Bio-Rad CFX 96 Touch RT-PCR system at the Curative Laboratories CLIA-certified laboratory in San Dimas, CA. All laboratories were blinded to specimen information and provided only the study ID. The Hologic Assay was chosen as the gold standard comparator due to its classification as a “high-sensitivity” assay and authorization for use in asymptomatic individuals by the FDA (5).

#### Institutional Review Board Oversight

The design of the study, including the protocol, recruitment materials, and consent forms, was approved by the Advarra Internal Review Board (IRB# PTL-2020-0003).

## Results

### Drive through Study

A total of 1,326 subjects ages 7 and older were enrolled, 971 (73.2%) were asymptomatic and 355 (26.8%) symptomatic. 1,274 (96.1%) subjects provided all swab types and 1,211 (91.3%) provided all swab types and a blood sample. Demographic information is provided in Supplementary Table S1.

The nasopharyngeal swab run on the EuroRT Assay was used as the comparator and Positive Percent Agreement (PPA) and Negative Percent Agreement (NPA) were calculated for each test, these data are summarised in Figure 1a & 1b and full diagnostics tables are provided in Supplementary Table S2.

**Figure 1.**
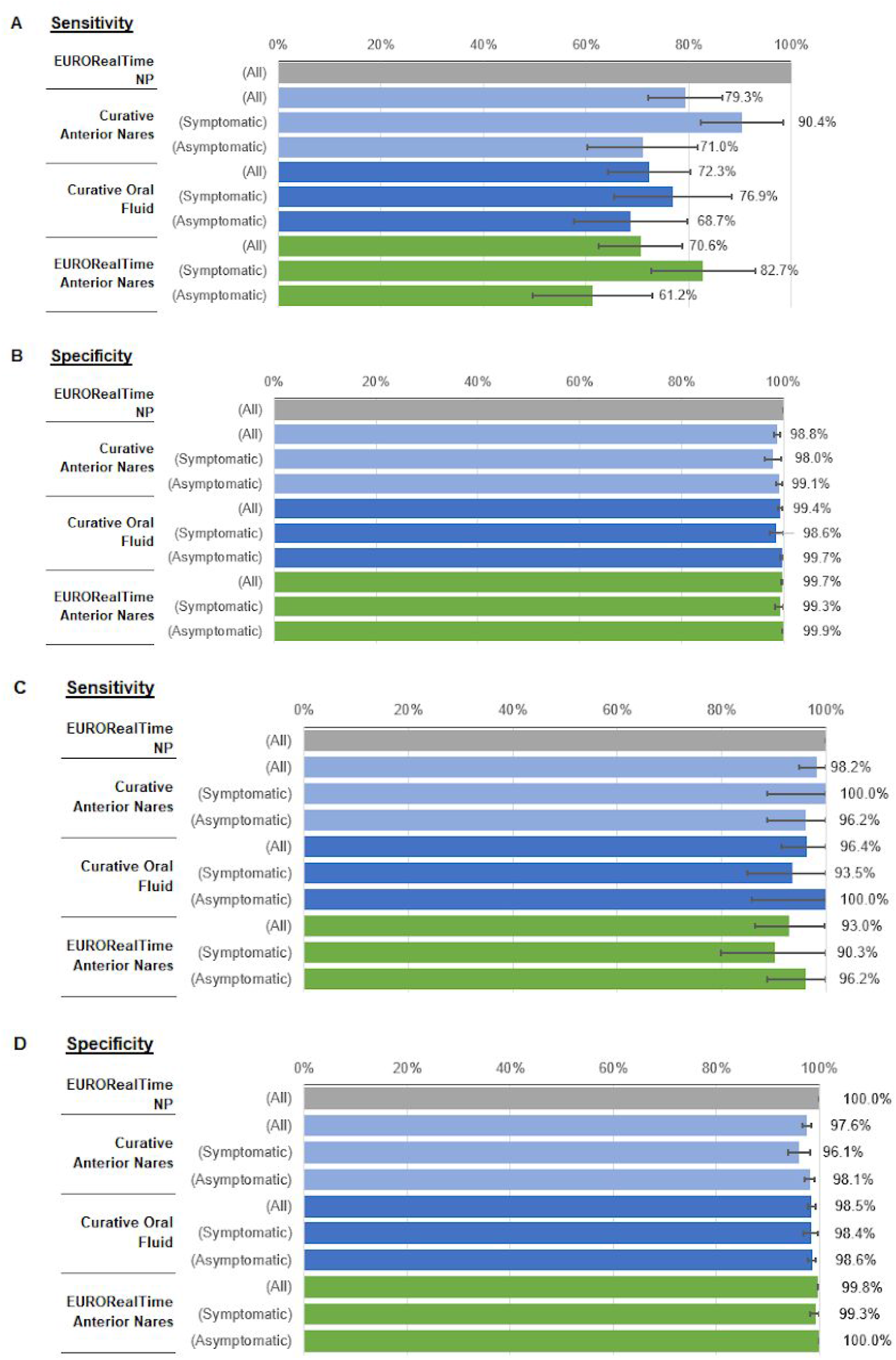
A & B Sensitivity & Specificity of Self-Collected Oral Fluid and Self-Collected Anterior Nares on the Curative SARS-CoV-2 Assay and Provider-collected Anterior Nares on the EURORealTime SARS-CoV-2 Assay compared a Nasopharyngeal Swab on the EURORealTime SARS-CoV-2 Assay as the reference, broken out by symptom status. C & D - The same sample comparison as in A & B but excluding NP RT-PCR positive subjects with a negative viral culture or positive IgG serology result

Self-collected anterior nares specimens run on the Curative Assay showed a PPA of 79.3% (95% Confidence Interval [CI]: 72.1% - 86.6%) and an NPA of 98.8% (95% CI: 98.2% - 99.4%). Self-collected oral fluid specimens on the Curative Assay showed a PPA of 72.3% (95% CI: 64.2% - 80.3%) and an NPA of 99.4% (95% CI: 99.0% - 99.8%). Interestingly when the provider-collected anterior nares specimen on the EuroRT Assay was compared against the provider-collected nasopharyngeal specimen run on the same RT-PCR Assay, the PPA was 70.6% (95% CI: 62.4% - 78.8%) and the NPA was 99.7% (95% CI: 99.5% - 100%).

### Correlation of RT-PCR, Viral Culture and Serology Results

PPA and NPA were then calculated with exclusion of individuals positive by NP RT-PCR but culture negative or serology positive, which would indicate these individuals do not have replication-competent virus and/or are late in disease based on the presence of anti-SARS-CoV-2 IgG (24) and therefore are not likely to be infectious. These data are summarised in Figure 1C & 1D and full diagnostic tables are provided in Supplementary Table S3.

When evaluating nasopharyngeal specimens from individuals that are more likely to be infectious as the reference, Curative anterior nares specimens showed a PPA of 98.2% (95% CI: 94.8% - 100.0%) and an NPA of 97.6% (95% CI: 96.7% - 98.5%). Curative oral fluid specimens showed a PPA of 96.2% (95% CI: 88.8% - 100.0%) and an NPA of 98.1% (95% CI: 97.2% - 99.0%). Provider-collected anterior nares specimen run on the EuroRT Assay showed a PPA of 93.0% (95% CI: 86.4% - 99.6%) and an NPA of 99.8% (95% CI: 99.6% - 100%). There was no significant difference between performance in asymptomatic or symptomatic individuals with PPAs in asymptomatic individuals of 96.2% (95% CI: 88.8% - 100.0%) and 100.0% (95% CI: 85.8% - 100.0%) in Curative anterior nares and oral fluid respectively.

### Early-COVID Results

All Curative Inc employees with a new positive SARS-CoV-2 test within the last 21 days were contacted, 61 consented and were enrolled in the study, of these 29 were asymptomatic and 32 were symptomatic at the time of sample collection. 60/61 subjects were confirmed positive by the reference test (Hologic Assay). Number of days from onset of infection was evenly distributed among subjects with a mean of 6.5 days and standard deviation of 6.3 days. 11 subjects were within 24 hours of onset of SARS-CoV-2 infection (determined as first positive SARS-CoV-2 test after prior daily negative tests) and 10 subjects were beyond 14 days of onset of SARS-CoV-2 infection. Subjects age ranged from 16 to 64 with a mean of 30

Additionally all Curative Inc. employees with no known history of SARS-CoV-2 infection and no positive SARS-CoV-2 test results were contacted, 104 consented and were enrolled in the study. All 104 individuals were asymptomatic and all were confirmed negative by the reference test.

The performance of the Curative SARS-CoV-2 Assay compared to the Hologic Aptima SARS-CoV-2 Assay in individuals within 21 days of onset of new SARS-CoV-2 infection is summarized in Table 1. A PPA of 100%, 100% and 98.3% and an NPA of 100%, 99.0% and 99.0% was seen for oral fluid, anterior nares and nasopharyngeal swabs respectively (confidence intervals listed in Table 1). There was no difference in performance between asymptomatic and symptomatic individuals and the Curative SARS-CoV-2 Assay showed 100% PPA and 100% NPA in all 3 specimen types in asymptomatic subjects.

**Table 1.** Comparison of Curative SARS-CoV-2 Assay with self-collected oral fluid, self-collected anterior nares and provider-collected NP specimens to Hologic Aptima SARS-CoV-2 with NP

| Curative SARS-CoV-2 Assay |  | Hologic Aptima with Nasopharyngeal Swab |  |  |  |
| --- | --- | --- | --- | --- | --- |
|  |  | All |  | Asymptomatic |  |
|  |  | Positive | Negative | Positive | Negative |
| Oral Fluid | Positive | 60 | 0 | 29 | 0 |
|  | Negative | 0 | 105 | 0 | 104 |
| Positive Percent Agreement (95% CI) |  | 100% (94.0% - 100%) |  | 100% (88.1% - 100.0%) |  |
| Negative Percent Agreement (95% CI) |  | 100% (96.5% - 100%) |  | 100% (96.5% - 100.0%) |  |
| Anterior Nares | Positive | 60 | 1 | 29 | 0 |
|  | Negative | 0 | 104 | 0 | 104 |
| Positive Percent Agreement (95% CI) |  | 100% (94.0% - 100%) |  | 100% (88.1% - 100.0%) |  |
| Negative Percent Agreement (95% CI) |  | 99.0% (94.8% - 100%) |  | 100% (96.5% - 100.0%) |  |
| Nasopharyngeal Swab | Positive | 59 | 1 | 29 | 0 |
|  | Negative | 1* | 104 | 0 | 104 |
| Positive Percent Agreement (95% CI) |  | 98.3% (91.1% - 100%) |  | 100% (88.1% - 100.0%) |  |
| Negative Percent Agreement (95% CI) |  | 99.0% (94.8% - 100%) |  | 100% (96.5% - 100.0%) |  |

\* This case was detected very early in the course of disease (day 0) and was positive by oral fluid and anterior nares but negative by NP on the Curative SARS-CoV-2 Assay

### Late-COVID Results

All Curative Inc. employees with a new positive SARS-CoV-2 result that occurred between 22 and 70 days prior to study recruitment were contacted. 22 subjects provided consent and were enrolled. 20/22 (90.9%) of subjects self-reported experiencing symptoms at some point during their SARS-CoV-2 infection. No subjects had been hospitalized during their SARS-CoV-2 infection. 3 subjects reported still experiencing mild symptoms at the point of sample collection. Number of days from onset of infection was evenly distributed among subjects from 28 to 64 days with a mean of 42.7 days and standard deviation of 10.4 days. Subjects age ranged from 20 to 50 with a mean of 30.

A much greater number of positive RT-PCR results were found with nasopharyngeal swabs compared to anterior nares or oral fluid swabs across all 4 EUA assays tested ranging from 4X to 18X more. Additionally, although Zymo Quick SARS-CoV-2 rRT-PCR Kit and Hologic Aptima SARS-CoV-2 Assay both detected a large number of NP RT-PCR positives (13 and 8 respectively), indicating a similar sensitivity, only 4 subjects were positive on both tests, 4 were negative on both and 14 results disagreed between the 2 tests. The positivity rates between sample types are shown in ***Figure 2*** and the results for each subject by sample type are summarized in ***Table 2***.

**Table 2.**
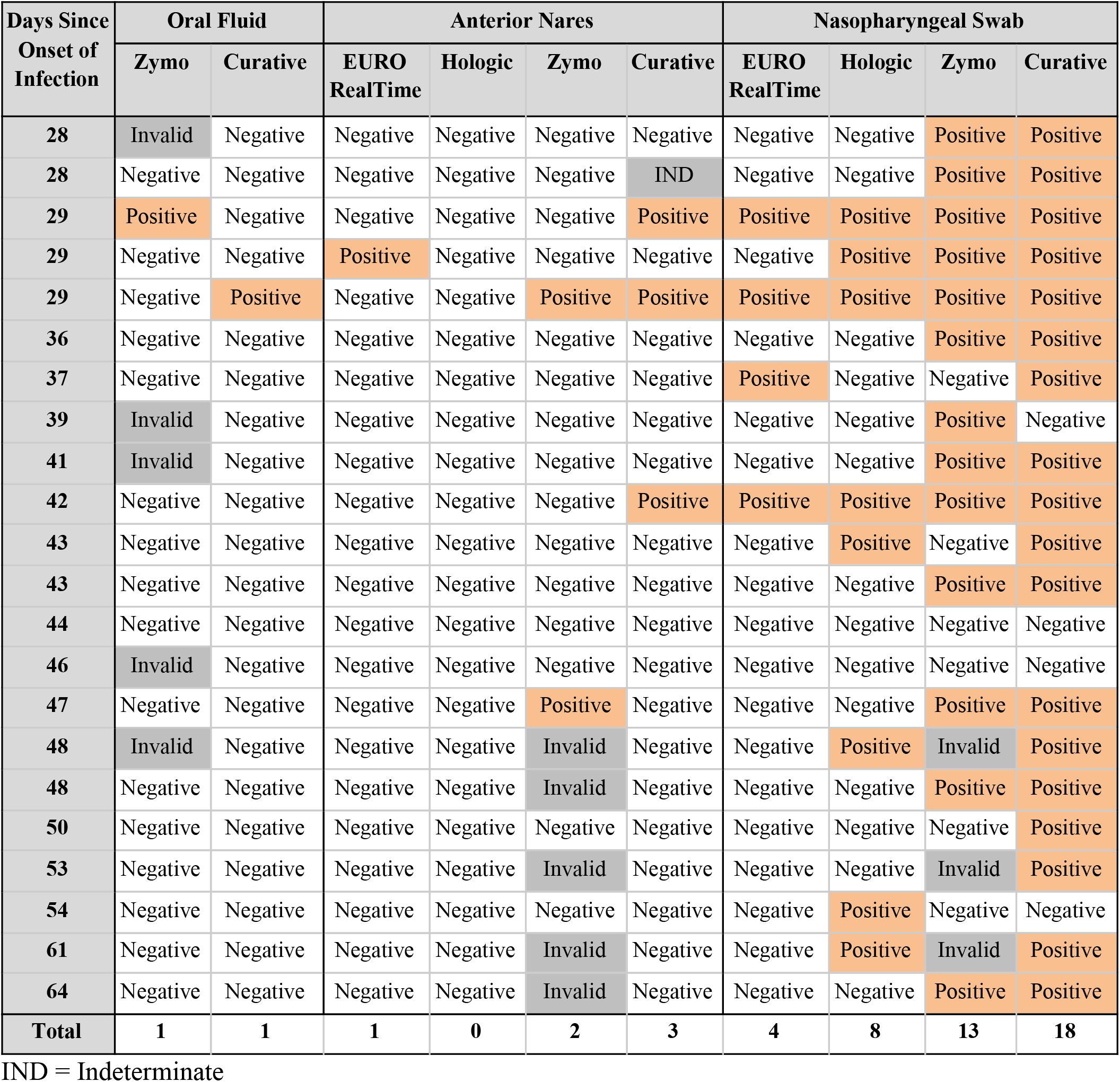
Comparison of incidence of positive test results at different types from the onset of SARS-CoV-2 infection by specimen type and assay

| Days Since Onset of Infection | Oral Fluid |  | Anterior Nares |  |  |  | Nasopharyngeal Swab |  |  |  |
| --- | --- | --- | --- | --- | --- | --- | --- | --- | --- | --- |
|  | Zymo | Curative | EURO RealTime | Hologic | Zymo | Curative | EURO RealTime | Hologic | Zymo | Curative |
| 28 | Invalid | Negative | Negative | Negative | Negative | Negative | Negative | Negative | Positive | Positive |
| 28 | Negative | Negative | Negative | Negative | Negative | IND | Negative | Negative | Positive | Positive |
| 29 | Positive | Negative | Negative | Negative | Negative | Positive | Positive | Positive | Positive | Positive |
| 29 | Negative | Negative | Positive | Negative | Negative | Negative | Negative | Positive | Positive | Positive |
| 29 | Negative | Positive | Negative | Negative | Positive | Positive | Positive | Positive | Positive | Positive |
| 36 | Negative | Negative | Negative | Negative | Negative | Negative | Negative | Negative | Positive | Positive |
| 37 | Negative | Negative | Negative | Negative | Negative | Negative | Positive | Negative | Negative | Positive |
| 39 | Invalid | Negative | Negative | Negative | Negative | Negative | Negative | Negative | Positive | Negative |
| 41 | Invalid | Negative | Negative | Negative | Negative | Negative | Negative | Negative | Positive | Positive |
| 42 | Negative | Negative | Negative | Negative | Negative | Positive | Positive | Positive | Positive | Positive |
| 43 | Negative | Negative | Negative | Negative | Negative | Negative | Negative | Positive | Negative | Positive |
| 43 | Negative | Negative | Negative | Negative | Negative | Negative | Negative | Negative | Positive | Positive |
| 44 | Negative | Negative | Negative | Negative | Negative | Negative | Negative | Negative | Negative | Negative |
| 46 | Invalid | Negative | Negative | Negative | Negative | Negative | Negative | Negative | Negative | Negative |
| 47 | Negative | Negative | Negative | Negative | Positive | Negative | Negative | Negative | Positive | Positive |
| 48 | Invalid | Negative | Negative | Negative | Invalid | Negative | Negative | Positive | Invalid | Positive |
| 48 | Negative | Negative | Negative | Negative | Invalid | Negative | Negative | Negative | Positive | Positive |
| 50 | Negative | Negative | Negative | Negative | Negative | Negative | Negative | Negative | Negative | Positive |
| 53 | Negative | Negative | Negative | Negative | Invalid | Negative | Negative | Negative | Invalid | Positive |
| 54 | Negative | Negative | Negative | Negative | Negative | Negative | Negative | Positive | Negative | Negative |
| 61 | Negative | Negative | Negative | Negative | Invalid | Negative | Negative | Positive | Invalid | Positive |
| 64 | Negative | Negative | Negative | Negative | Invalid | Negative | Negative | Negative | Positive | Positive |
| **Total** | **1** | **1** | **1** | **0** | **2** | **3** | **4** | **8** | **13** | **18** |

IND = Indeterminate

**Figure 2.**
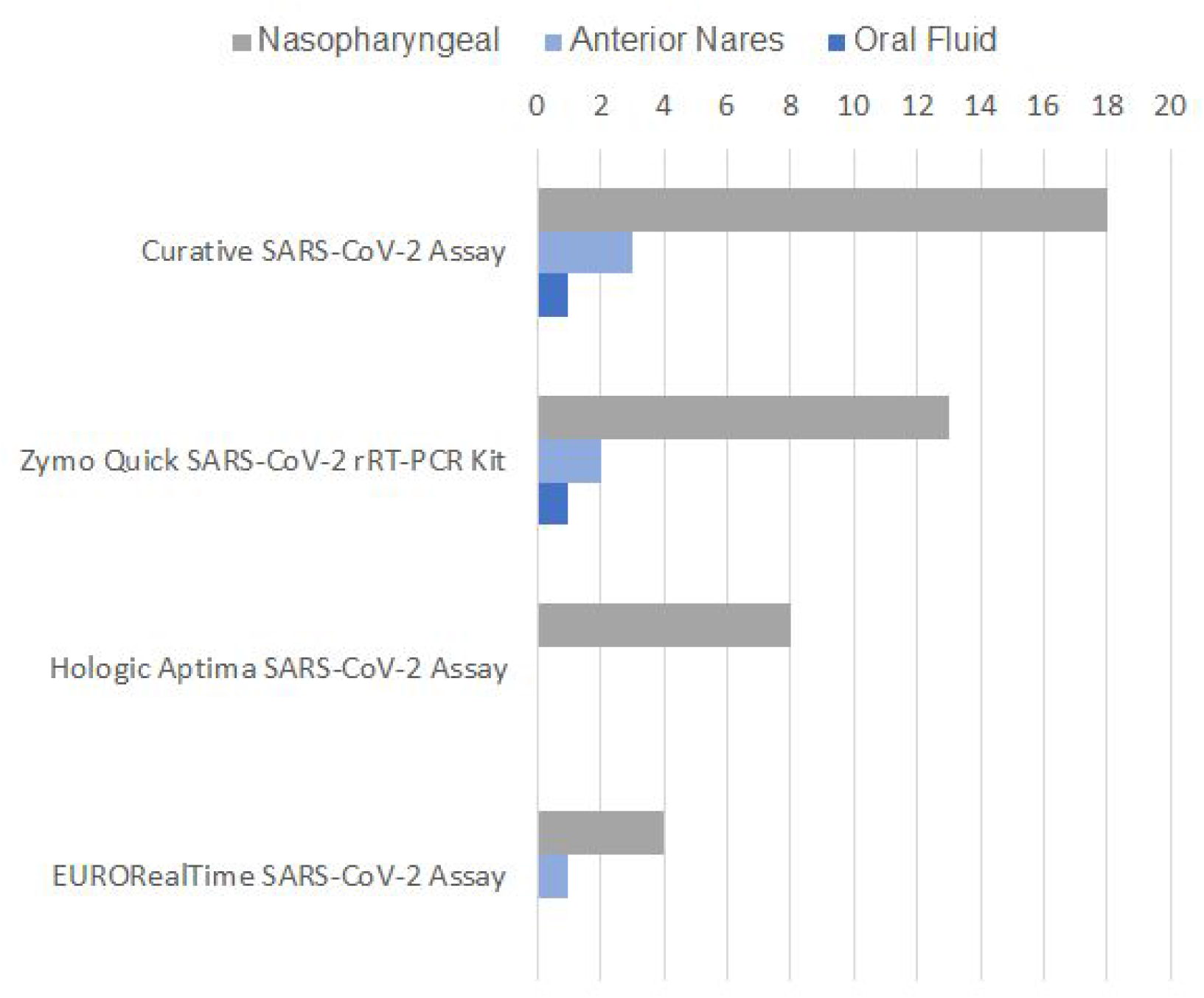
Number of positive molecular test results by specimen type on each of the 4 EUA tests in the late-COVID study (subjects > 21 days from onset of infection)

## Discussion

### Real World “Drive-through” Specimen Comparison Study

The COVID-19 pandemic has driven a massive expansion in diagnostic testing, to facilitate this while following social distancing guidelines, much of this testing is being conducted in non-traditional clinical settings such as drive-throughs, which can test over 10,000 patients in a single day (Curative Internal Data). These testing sites have highly complex patient mixes with limited medical history. While these populations typically encompasses 50-60% of patients testing due to symptoms or exposure, many patients also test regularly for employment or travel with 51.4% of patients testing multiple times including 13.4%, 6.8% and 0.5% who self-reported testing monthly, weekly or daily, respectively (Curative Internal Data, n=241,986). Hence, many patients are tested late in their course of clinical disease or may knowingly test again within 3 months of a resolved SARS-CoV-2 infection, despite CDC guidelines to the contrary. (26).

We compared self-collected oral fluid and anterior nares samples on the Curative SARS-CoV-2 Assay to provider-collected NP and anterior nares samples on the EuroRT Assay in two “drive-through” testing sites. Taking the NP swab on the EuroRT Assay to be the reference standard we observe a similar sensitivity between the two anterior nares and oral fluid specimens, with PPAs of 79.3%, 72.3% and 70.6% for Curative anterior nares, Curative oral fluid and EuroRT anterior nares, respectively (***Fig 1A***). However, 20-30% more positive results were detected with the NP specimen. It is noteworthy that the PPA of anterior nares compared to NP is only 70.6% when using the same RT-PCR assay and sample collection system (EuroRT Assay). This strongly suggests the observed difference is due to biological variation between the specimens. Unfortunately, an NP sample for evaluation using the Curative Assay was not collected in the initial drive through study. Hence, given the apparent biological variation between specimen types, evaluating the performance of a different RT-PCR assay while also varying the specimen type is very challenging.

We anticipated this specimen variation and included viral culture and serology testing in the drive through study design. If subjects that are only positive by NP RT-PCR are late in the course of disease, we would expect the NP RT-PCR Cycle Threshold (Ct) to be high, no replication-competent virus to be recoverable through viral culture testing, and the subjects to have detectable anti-SARS-CoV-2 IgG-class antibodies.

Of the subjects positive by NP RT-PCR with viral culture data available, 30 were negative on oral fluid and 23 negative on anterior nares RT-PCR on the Curative Assay and 32 were negative on anterior nares RT-PCR on the EuroRT Assay (See ***Figure 3***). The mean Cts on the NP RT-PCR were 35.04 and 35.26 for subjects with negative oral fluid and anterior nares RT-PCRs respectively.

**Figure 3.**
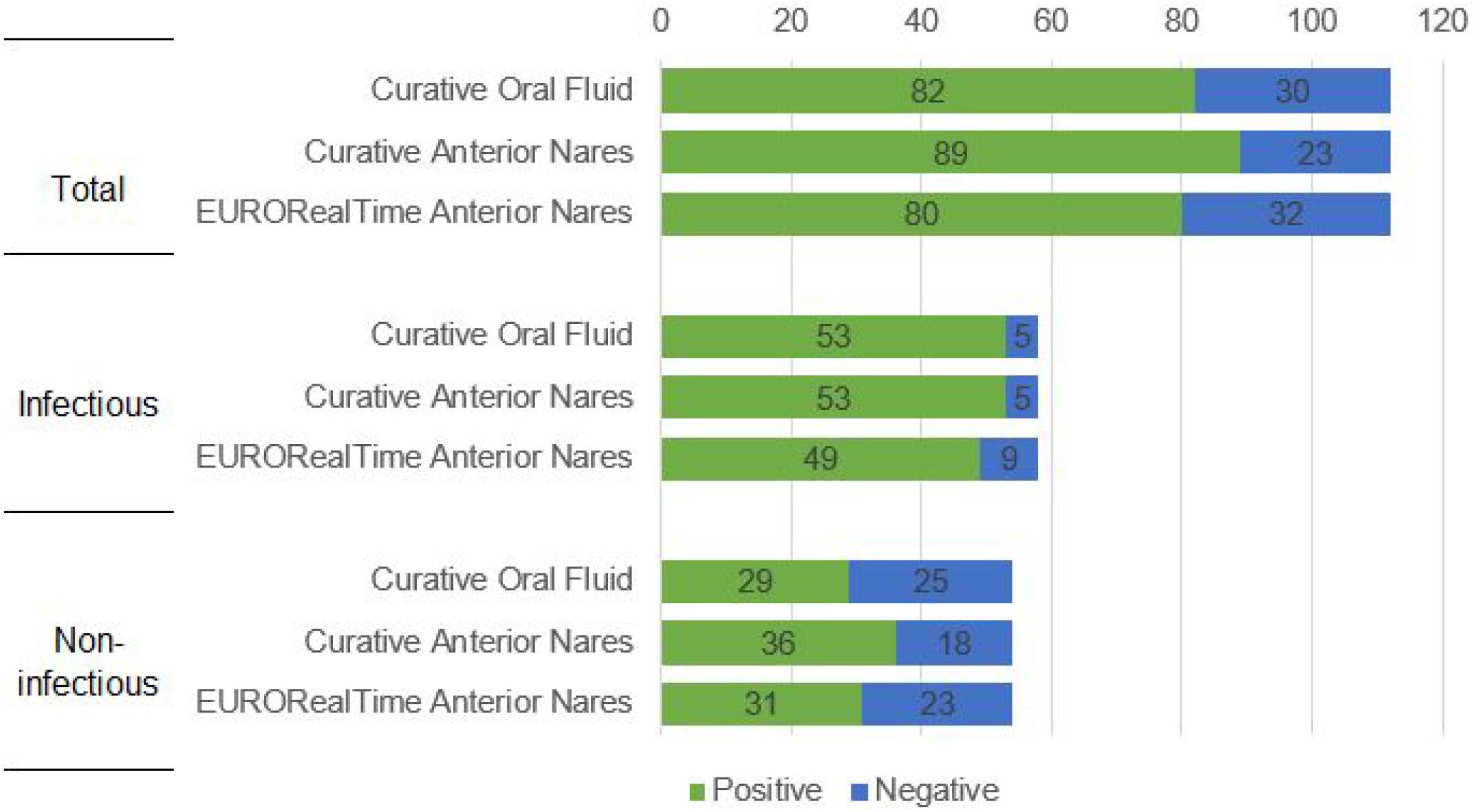
Break down of subjects with a positive NP specimen by EURORealTime RT-PCR. The RT-PCR test result of the other paired samples is shown in the bars and the subjects are further split out into those with a positive viral culture result (Infectious) or a negative viral culture result (Non-infectious). Five subjects without a viral culture result (Test Not Performed) were excluded.

Of the 58 subjects with positive NP RT-PCR that were also positive by viral culture 53/58 (91.4%) were positive by oral fluid and anterior nares on the Curative Assay compared to 49/58 (84.5%) with anterior nares on the EuroRT Assay (***Figure 3***)

In our study we observed 12.1 % of the tested population had previously been infected with SARS-CoV-2 based on the detection of IgG which is produced on average 14 days after infection (24). The presence of a large post-Covid cohort may impact current testing strategies depending on the specimen type used due to the persistent-positivity on NP swab specimens significantly inflating test positivity rates and overestimating the number of active infections.

Looking at NP positivity rates in absence of infectious status suggests that oral fluid and anterior nares specimens, regardless of assay, showed a lower sensitivity than the NP comparator. One explanation of these data is that recruitment of individuals at drive through testing sites with a high RT-PCR positivity and seroprevalence of SARS-CoV-2, is likely to enroll many individuals previously infected with SARS-CoV-2 who are now recovered but remain persistently positive in NP specimens and an accurate assessment of other specimen types cannot be made without adjusting for stage of infection using viral culture and serology data. An alternative explanation is that the sensitivity of the Curative Assay is insufficient to detect these low viral load positives. This second explanation is not supported by the performance of the anterior nares specimen on the EuroRT Assay mirroring that of the Curative assay however we determined to definitely prove this is truly biological variation between NP and other specimen types late in clinical disease using a population with a highly characterized date of onset of infection.

### Separating Assay and Specimen Variability Over the Time Course of SARS-CoV-2 Infection

Shedding of SARS-CoV-2 viral RNA in NP swab specimens long after clinical disease is well documented in the literature (1, 2, 17, 18) however this process and its cause is poorly understood. To test the hypothesis that this persistent-positivity after resolution of clinical disease occurs to a significantly greater extent with NP swab specimens than anterior nares or oral fluid, we utilized a unique population with a comprehensive history of daily SARS-CoV-2 testing, allowing precise determination of the onset of new infection.

In this population we showed comprehensively that during active SARS-CoV-2 infection (< 21 days from infection onset) oral fluid, anterior nares and nasopharyngeal specimens show a high degree of consistency when compared either with the same assay or between different assays and showed PPAs of 98-100% and NPAs of 99-100% for all specimen types on the Curative SARS-CoV-2 Assay when compared to NP swabs run on a high-sensitivity comparator assay (Hologic Aptima SARS-CoV-2 Assay) (See ***Table 1***). The data further confirms, as with other molecular tests, that there is no difference in test performance between symptomatic and asymptomatic individuals and the performance of the Curative SARS-CoV-2 is equivalent to other high-sensitivity molecular assays.

Further, we demonstrated a transition occurring between 22 and 28 days after onset of infection where oral fluid and anterior nares viral loads consistently drop to zero while nasopharyngeal viral RNA persists for the full duration measured (up to 64 days) (***Figure 4*** shows the full time course of infection compared between specimen types). We showed that this effect is observed with 4 different FDA EUA molecular tests including the Hologic Aptima SARS-CoV-2 Assay which is a high-sensitivity assay explicitly authorized for testing of asymptomatic individuals (5). Thus, we clearly demonstrate that this effect is due to biological variation between specimen types and not due to the assay used for detection.

**Figure 4.**
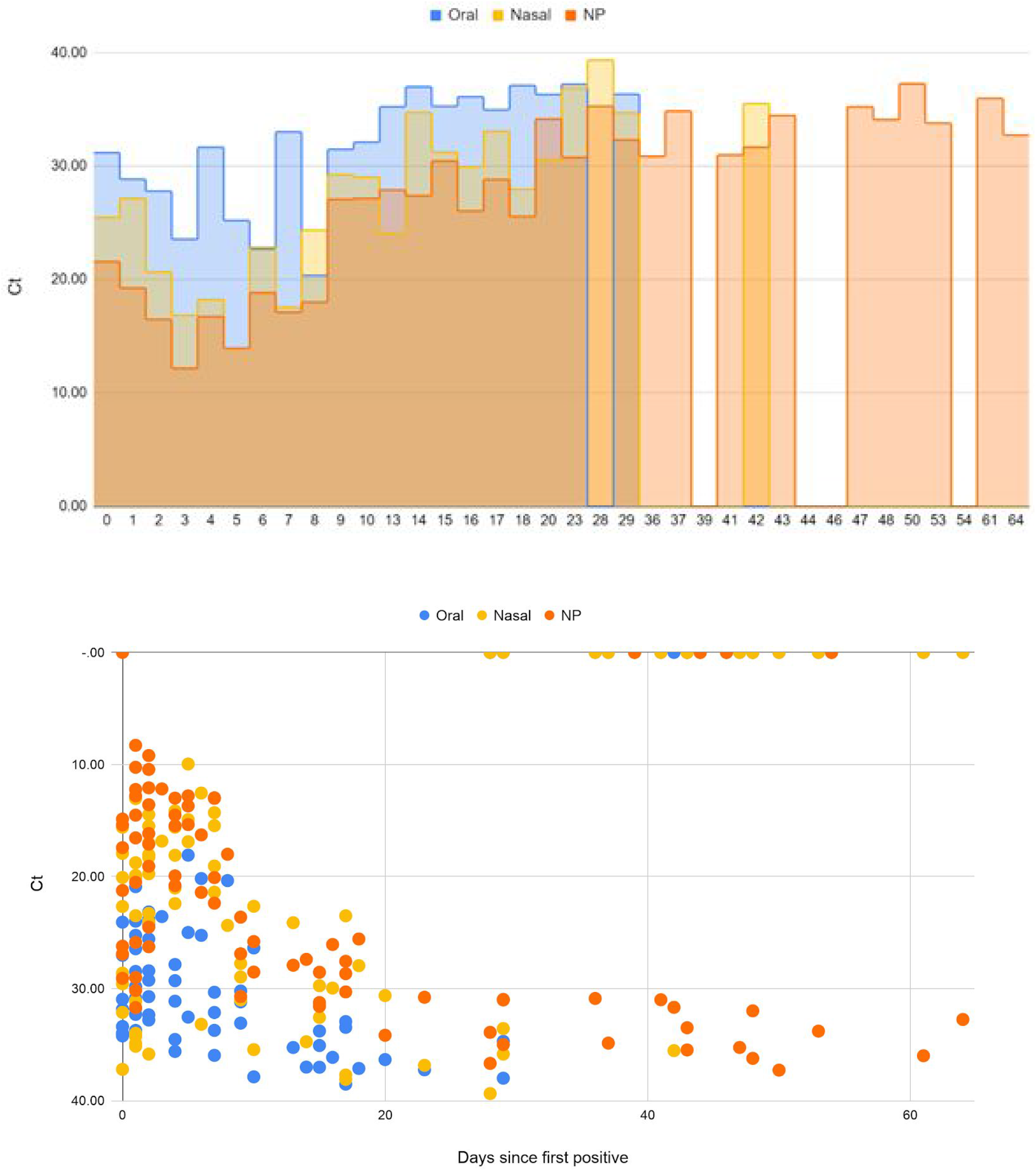
Upper Panel: Mean Ct for subjects by days since first positive test, broken out by specimen type. Lower Panel: Scatter plot of the same with all individual Cts. Zero values represent a negative test results

### Biological mechanisms

We observed varying levels of detection of persistent-positives on NP swabs, late in disease, between different assays used to detect them, ranging from 18/22 to 4/22 specimens testing positive. Possibly, the amount of viral RNA is so low that stochastic sampling bias is introduced. The Curative and Zymo assays, which detected 18/22 and 13/22 positives respectively, both utilize DNA/RNA Shield for specimen collection, a guanidinium-based preservative that deactivates nucleases and preserves viral RNA. Whereas, the Hologic and EuroRT Assays, which detected 8/22 and 4/22 positives respectively, utilize Viral Transport Media (VTM), that does not deactivate nucleases or actively preserve RNA stability. This data may indicate that DNA/RNA Shield improved the sensitivity of RT-PCR tests.

The Curative and Zymo assays both target two and three segments of the N (nucleocapsid) gene respectively (3, 6), the Hologic Aptima Assay targets two segments of ORF1ab (5) and the EuroRT Assay targets one segment of ORF1ab and one in the N gene (4). During viral replication, sub-genomic RNAs (sgRNAs) are generated and discontinuous transcription is expected to produce a significantly greater quantity of sgRNAs containing the N gene than ORF1ab (26, 27), possibly accounting for the increased rate of detection in assays exclusively targeting the N gene.

The mechanism of long term RNA persistence is not currently understood. Attempts to isolate replication-competent virus beyond 20 days have repeatedly failed (19, 20, 21, 22) and no mechanism for viral particles or free-RNA to persist extracellularly in the nasopharynx for over 30 days is readily apparent. One possible explanation is the poorly understood process of SARS-CoV-2 viral replication which involves viral proteins and RNA being sequestered in double-membrane vesicles in the cytoplasm and extensive remodeling of intracellular membrane systems (29). Such vesicles have been shown to be nuclease resistant and may persist intracellularly after active viral infection (30). This RNA would then ultimately get released upon cell death and hence the differential presence of remnant viral RNA between the nasopharynx and the anterior nares/oral cavity can be explained by the reduced percentage of vulnerable ACE2 expressing cells in these locations (31), the significantly slower rate of epithelial cell turnover in the nasopharyngeal mucosa compared to the buccal or anterior nasal mucosa as well as the greater abrasive forces applied to the epithelium during collection of a nasopharyngeal swab than an anterior nares or oral fluid swab which are likely to release more intracellular material. Further investigation into possible mechanisms is warranted.

### Public Health Implications - Nasopharyngeal swabs should not be used at community testing sites

Current CDC guidelines recommend discontinuing isolation either 10 days after symptom onset or 10 days after the first positive RT-PCR test for persons who never develop symptoms (23). This is based on substantial evidence that beyond this point, replication-competent virus cannot be isolated and repeated studies have shown individuals are not likely to be infectious (19, 20, 21, 22). Persistent viral RNA shedding in recovered individuals is well known and without new symptoms CDC do not recommend retesting within 90-days of recovering from SARS-CoV-2 infection, even with close contact with an infected person (26).

However, these guidelines are frequently ignored by individuals using community testing sites to obtain a negative test result required for employment, travel, etc. Moreover, CDC guidelines are omitted from EUA Summaries, Patient and Provider Factsheets provided by FDA for the interpretation of positive RT-PCR test results.

Differences in viral RNA shedding between specimen types are rarely considered in the choice of specimen type at community testing sites, however as the pandemic progresses, a substantial proportion of the population at these sites may have recovered from SARS-CoV-2 infection in the past 90 days. In this setting the Positive Predictive Value (PPV) of NP specimens, for clinical disease where isolation or treatment are beneficial, is substantially reduced and may be less than 50%. Isolating and contact tracing individuals late in disease is counterproductive and utilizes scarce resources with little or no public health benefit whilst also removing individuals from the workforce unnecessarily during a time of significant economic upheaval.

In this setting anterior nares or oral fluid specimens are likely to provide better PPV of acute SARS-CoV-2 infection and focus resources on individuals where a public health benefit can be derived from acting on the test results. Re-evaluating testing programs in light of these new findings is imperative to ensure resources are allocated appropriately.

### Policy Implications - Nasopharyngeal swabs

These findings also suggest that nasopharyngeal swabs are an inappropriate comparator for the evaluation of novel assays and sample types. The FDA has moved rapidly to authorize hundreds of molecular tests for Emergency Use during the COVID-19 pandemic and has dramatically expanded access to testing (32). However, the number of tests authorized for alternative specimen types such as oral fluid or saliva which can provide easy, high-throughput access to regular testing, has remained small (33), despite these specimen types being shown to be highly effective (15, 34, 35, 36, 37). And in fact there still remains no test authorized for use in asymptomatic screening using saliva or oral fluid. Similarly the number of antigen tests, which could provide rapid, low-cost identification of positive cases has remained low.

Our findings suggest this may be more due to the use of NP swabs as the comparator than to the performance of these novel tests or specimen types. Both novel RT-PCR specimen types and antigen tests provide a more accurate assessment of infectious potential than NP RT-PCR and are advantageous to public health. However, current FDA templates for Molecular Diagnostic Developers require NP swabs as the comparator for new specimen types (38). It is clear that when evaluated in a study population with subjects who have recovered from SARS-CoV-2 infection in the last 90 days, the sensitivity of these tests is likely to appear artificially low, because the variable being measured is the proportion of persistent positive viral RNA shedders in the study population and not the sensitivity of the assay or specimen in question.

This effect is particularly pronounced in the evaluation of tests in asymptomatic individuals where finding a study population with the required 20 asymptomatic positive cases (38) but no individuals, currently without symptoms, who have recovered from SARS-CoV-2 infection > 21 days prior, is almost impossible. The extreme example of following thousands of known SARS-CoV-2 negative individuals for months with daily testing to identify asymptomatic infections without identifying individuals previously infected > 21 days prior, while effective for demonstrating that in an appropriate study population, oral fluid is highly-sensitive for detecting asymptomatic SARS-CoV-2 infection, is impractical for the majority of test developers. Hence, current policy significantly restricts the availability of tests that may be highly effective in preventing the spread of SARS-CoV-2.

Multiple groups have reported widely variable results in comparing saliva/oral fluid to NP swabs (good: 15, 37, 39, 41, 45 bad: 40, 42, 43, 44) including some reports of variability with the same assay between study locations (40). The source of this variability has not been fully understood but we suggest that variability in the proportion of people with SARS-CoV-2 infection >21 days prior is the likely root cause. More extensive use of saliva and oral fluid specimen types with new and existing assays could be highly beneficial to expand access to testing. For this to occur it is vital that test developers understand the implications of the study population within which they choose to evaluate their assay and that regulatory policy evolves to use a more clinically appropriate comparator that will provide consistent results between studies.

### Limitations of Studies

The studies presented here were conducted predominantly in groups of mild to moderate severity COVID-19 cases and no hospitalized patients were included in the group. Although many studies recruited in academic medical centers for SARS-CoV-2 have introduced the opposite bias and understanding, testing and disease progression in an outpatient setting is vital for managing the SARS-CoV-2 pandemic, there may be differences in the effects described here in severe COVID-19 cases requiring treatment and hospitalization as they were not included in the present study. Additionally the extensibility of these data to children under the age of 16 is unknown.

Previous surveillance test data, which is performed with either oral fluid or anterior nares specimens on the Curative Assay, was used to recruit suspected positive subjects to the study. The possibility of a sampling bias here is reduced by the high frequency at which this employee surveillance testing was conducted (daily or twice-daily) and by the large suspected negative group which was recruited from the same population using the same criteria, except without the recent positive test result. The current prevalence of SARS-CoV-2 in Los Angeles, CA, Austin, TX and Washington, DC, where these studies were conducted, is very high and in the eligible study population is ∼3-4%. Therefore, if true positive cases were being missed by the surveillance testing that would have been caught on the reference test, we would expect multiple positive cases to be identified by the reference test in the suspected negative group and this was not the case.

## Data Availability

All raw data available upon request.

## Notes

### Competing Interest Statement

Fred Turner,  Amy Vandenberg, Vladimir Slepnev, Suzana Car, Nina Nirema, Lauren Lopez, Matthew Brobeck, Sarah August, Alejandra Orosco, Fred Hertlein and Arthur Baca are employees of Curative Inc. which funded the study and distributes the Curative SARS-CoV-2 Assay studied.
Fred Turner and Vladimir Slepnev are shareholders in Curative Inc.

### Clinical Trial

Non-interventional specimen collection trials only. No NIH/US Gov funding received.

### Funding Statement

All work was funded by Curative Inc.

### Author Declarations

The design of the study, including the protocol, recruitment materials, and consent forms, was approved by the Advarra Institutional Review Board (IRB# PTL-2020-0003).

